# A text messaging intervention to support patients with chronic pain during prescription opioid tapering: protocol for a double-blind randomised controlled trial

**DOI:** 10.1101/2023.05.08.23289534

**Authors:** Ali Gholamrezaei, Michael R Magee, Amy G McNeilage, Leah Dwyer, Hassan Jafari, Alison Sim, Manuela L Ferreira, Beth D Darnall, Paul Glare, Claire E Ashton-James

## Abstract

**Introduction:** Increases in pain and interference with quality of life is a common concern among people with chronic non-cancer pain (CNCP) who are tapering opioid medications. Research indicates that access to social and psychological support for pain self-management may help people to reduce their opioid dose without increasing pain and interference. This study evaluates the efficacy of a text messaging intervention designed to provide people with CNCP with social and psychological support for pain self-management while tapering long-term opioid therapy (LTOT) under the guidance of their prescriber.

**Methods and analysis:** A double-blind randomised controlled trial will be conducted. Patients with CNCP (*n* = 220) who are tapering LTOT will be enrolled from across Australia. Participants will continue with their usual care while tapering LTOT under the supervision of their prescribing physician. They will randomly receive either a psycho-educational video and supportive text messaging (two SMS per day) for 12 weeks or the video only. The primary outcome is the pain intensity and interference assessed by the Pain, Enjoyment of Life and General Activity scale. Secondary outcomes include mood, self-efficacy, pain cognitions, opioid dose reduction, withdrawal symptoms, and acceptability, feasibility, and safety of the intervention. Participants will complete questionnaires at baseline and then every four weeks for 12 weeks and will be interviewed at week 12. This trial will provide evidence for the efficacy of a text messaging intervention to support patients with CNCP who are tapering LTOT. If proven to be efficacious and safe, this low-cost intervention can be implemented at scale.

**Ethics and dissemination:** The study protocol was reviewed and approved by the Northern Sydney Local Health District (Australia). Study results will be published in peer-reviewed journals and presented at scientific and professional meetings.

**Trial registration number:** ACTRN12622001423707.

**Strengths and limitations of this study:**

1. This protocol describes a double-blind randomised controlled trial in which patients, clinicians, investigators, outcome assessors, and statistician will all be blind to group allocation.
2. The intervention was co-designed with input from both patients and clinicians and further refined based on feedback received during a pilot study of the intervention.
3. The design and procedures of this trial were informed by the feasibility and potential efficacy measures of a pilot trial.
4. By using a broad community-based sample, this study aims to conduct more inclusive pain research and investigate the barriers and facilitators of implementing digital support on a larger scale.
5. Designing a sham or placebo control that is completely identical to the intervention is a challenge for a digital intervention. The active control may provide benefits to its recipients, potentially leading to an underestimation of the actual effect size of the digital support.

## Introduction

### Background and rationale

Chronic pain conditions are among the leading causes of disability worldwide.^1^ Whilst opioid medications are commonly prescribed for the management of chronic non-cancer pain (CNCP),^2^ prescribing guidelines have changed recently due to evidence of limited benefits and dose-related harms associated with long-term opioid therapy (LTOT).^3^ Consequently, patients with CNCP are increasingly being advised to gradually reduce the dose or discontinue LTOT (i.e., tapering) under clinical supervision.^4^

Systematic reviews suggest that, on average and over the long term, tapering opioids for patients with CNCP is not associated with increased pain and can be associated with improved functioning and quality of life.^5 6^ However, patients’ experiences of opioid tapering vary widely.^7^ Opioid dose reduction can lead to unpleasant withdrawal symptoms and negatively impact mood and pain.^7 8^ Recent retrospective studies have also raised concerns about the risk of overdose and serious harm associated with opioid tapering.^9^ Several studies indicate that the trajectory of patients’ experiences during tapering LTOT may depend on their access to various forms of support, including pain education, routine monitoring, a strong patient-provider relationship, and strategies for managing pain and withdrawal symptoms.^5 7 10 11^ However, access to these forms of support is often a pervasive challenge.^8 12 13^

Digital health technologies using mobile phones (mobile health or mHealth) are emerging as a solution to the global challenge of providing patients with access to support for health behaviour change.^14 15^ They can be a cost-effective way to deliver healthcare services on a large scale, particularly when they are used to address chronic conditions, and can be readily adapted to the needs of diverse demographic groups and health conditions.^14 16 17^ A recent systematic review found that digital health interventions can help to improve pain interference and severity, psychological distress, and health-related quality of life in people with chronic pain.^18^ However, evidence for the effectiveness of digital health interventions to support patients with CNCP during tapering LTOT is limited but promising.^19^

Our previous research has shown that people with CNCP have generally positive attitudes towards using digital health technologies, particularly mobile text messaging (SMS), to support them with opioid tapering.^20^ Studies have also shown that educational videos may be an effective means of providing patients with information about chronic pain, pain self-management, and opioid tapering,^21-23^ and can increase the opioid-tapering self-efficacy of people currently on LTOT for chronic pain.^21^ Based on this foundational research, we co-designed a digital intervention (video and mobile text messaging) for patients with CNCP who are tapering prescription opioids under clinical supervision.^24^ Patients and clinicians have rated this program as appropriate, useful, and likely to be effective in supporting patients during opioid tapering.^24^ A pilot randomised controlled trial (RCT) has been conducted, demonstrating the feasibility, acceptability, and potential efficacy of this intervention in patients referring to public and private outpatient pain management clinics.^25 26^

Therefore, in this study, we aim to evaluate the efficacy (primary objective) of our text messaging intervention to improve patients’ outcomes in a larger and more diverse population in Australia. To conduct more inclusive pain research,^27^ this study will use a broad community sample rather than merely recruiting through specialist pain clinics which are not easily accessible to most patients.^28^ Using a community sample will also enable us to investigate the barriers and facilitators of implementing this digital support on a large scale and to prepare strategies accordingly. Since, compared to our pilot RCT, the study setting and population have changed in this RCT and some aspects of the intervention have been modified (e.g., duration and content), we will re-evaluate the feasibility, acceptability, and safety of the intervention (secondary objectives). If found to be feasible, acceptable, safe, and efficacious, this digital health support can be evaluated in future cost-effectiveness/implementation trials as a solution to improve outcomes and reduce the harms of LTOT in patients with CNCP.

## Objectives

The primary objective of this RCT is to evaluate the efficacy of a text messaging intervention in reducing pain intensity and interference (as measured by the Pain, Enjoyment of Life and General Activity (PEG) scale)^29^ in patients with CNCP during tapering LTOT. Secondary objectives are A) to evaluate the efficacy of the intervention in reducing the severity of anxiety and depression symptoms, increasing opioid tapering self-efficacy and pain self-efficacy, reducing pain catastrophising, reducing the opioid dose, and reducing withdrawal symptoms, B) to evaluate the feasibility, acceptability, and safety of the intervention, C) to explore mechanisms of action and predictors of response to the intervention, and D) to identify barriers and facilitators of future implementation of the intervention at large-scale.

## Methods and analysis

This report is prepared according to the Standard Protocol Items: Recommendations for Interventional Trials (SPIRIT, see Appendix 1 for the SPIRIT checklist).

### Patient and public involvement

The development of this mHealth intervention was in response to previous research conducted by our team, as well as others, on patients’ experiences and needs during tapering opioid medications,^7 8 12^ and their perspectives on receiving support via mHealth interventions.^20^ As described previously, the mHealth intervention prototype was developed in close collaboration with clinician-researchers and a consumer representative.^24^ It was revised in response to feedback from panels of consumers and clinical experts, and then was tested in a pilot study.^25^ The intervention content and delivery were further improved based on the feedback received from those who participated in the pilot study.^26^ Similarly, the design (procedures and measures) of the pilot trial^25^ as well as the current, more definitive trial was developed in close collaboration with clinician-researchers and a consumer representative. Moreover, feasibility data and feedback received from those who participated in the pilot study were considered in designing this trial.^26^

### Study design

This is a double-blind RCT with two parallel arms (Figure 1) to test the primary hypothesis which is the superiority of receiving an educational video followed by daily text messages (SMS) compared with receiving an educational video alone, both in addition to usual care, in reducing pain intensity/interference.

**Figure 1:**
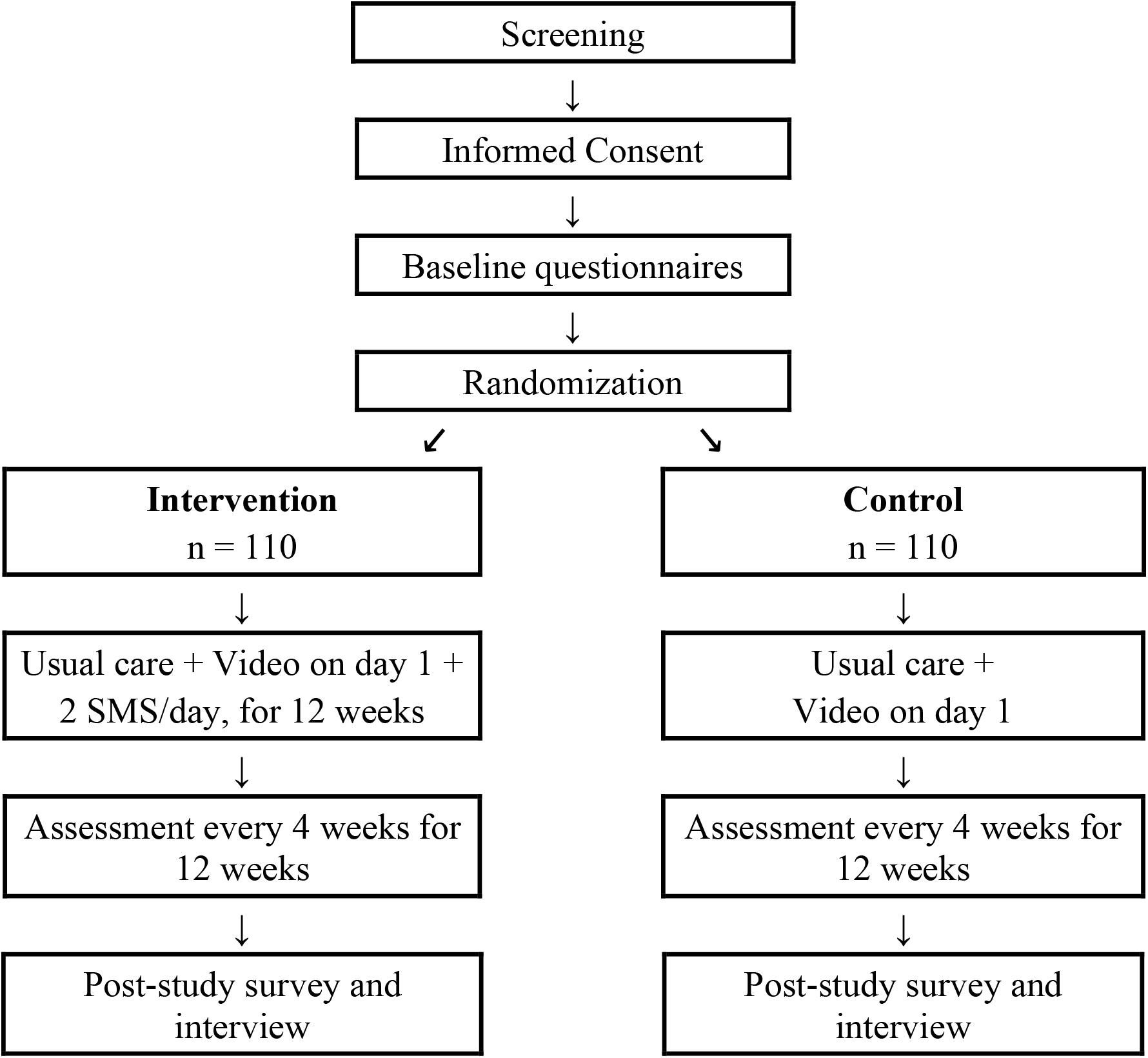
Study Flow Diagram.

There is currently no gold-standard ‘placebo’ control for digital health interventions and designing a sham condition for such interventions is challenging. Using an active control condition (i.e., educational video) together with ‘limited disclosure’ in the current study (see *Blinding/Masking*) will allow us to achieve blinding (masking) while complying with the national standards on ethical conduct in human research.^30^

### Study setting

Participants will be recruited from across Australia. Flyers (Appendix 2) will be used to identify potential participants via social media, newspapers, public notice boards, community events, websites, and email lists.

### Participants, screening, and recruitment procedures

The study flyers contain a link to the study webpage (Appendix 2) which provides brief information about the study and access to the Participant Information Sheet (PIS, Appendix 3) and an online self-report screening form (Appendix 4) to check eligibility (Table 1). Those who are eligible will be requested to consent online (Appendix 3).

**Table 1:**
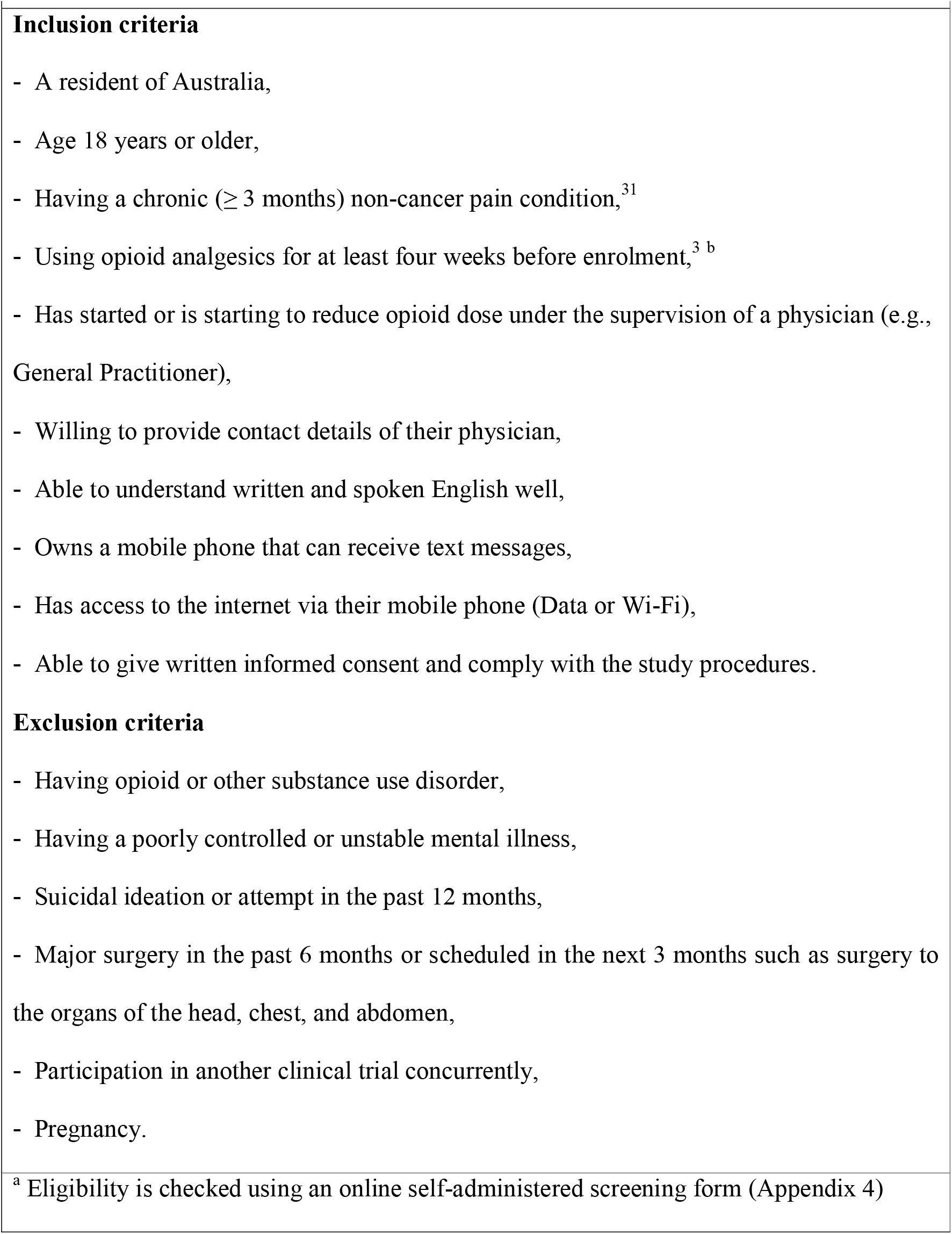
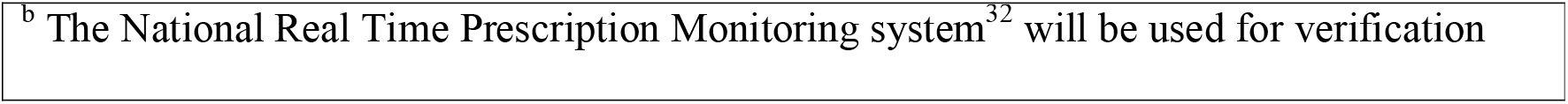
Eligibility criteria ^a^

### Interventions

#### Usual care

In this study, usual care is defined as any care that each participant receives at the discretion of their prescriber (e.g., their General Practitioner), which may or may not include multidisciplinary care. Participants in the current study are those who are tapering their opioid medications under the supervision of their prescribing physician. This physician is responsible for the monitoring and management of withdrawal symptoms and other possible adverse events during opioid tapering. Importantly, participation in this clinical trial does not involve clinical decisions around reducing opioids, the rate of dose reduction, or any other care that participants may be receiving. Participation in this study lasts for a total duration of 12 weeks.

#### Intervention group

After signing the consent form and completing baseline questionnaires, participants will be randomised (1:1) into two groups (Figure 1). Participants in the intervention group will receive a welcoming message and a message with a link to a psycho-educational video to watch (day 1) followed by daily text messages on their mobile phones for 12 weeks. The video and library of the messages were developed based on a co-design study, driven by the Social Cognitive Theory (with emphasis on the self-efficacy construct) and direct feedback from consumers and clinicians.^24^ The contents have been revised further based on feedback received during a pilot RCT.^25^ The psycho-educational video is composed of narrated, animated PowerPoint slides with a duration of 11 minutes providing information about pain, opioid tapering, and pain self-management strategies as well as socioemotional support in the form of testimonials. The content of the text messages serves to reinforce and remind people about the content of the psycho-educational video as well as messages of support and reassurance (see examples in Table 2 and Appendix 5).^25^ Starting from day 2, participants will receive daily text messages (2 SMS/day, 7 days/week) for 12 weeks (135 unique messages, 20% repeated messages, and 5 messages for assessment notifications). The text messaging duration for this study is based on patients’ feedback in the pilot study.^25^

**Table 2:** Examples of SMS text messages

| **Table 2: Examples of SMS text messages** |
| --- |
| **Pain management information**<br>*“Pain flare ups are like bad moods. Sometimes they come on for no clear reason. They will often pass if you distract yourself. Try doing or thinking about something you enjoy.”*<br>*“Slow, deep breathing can help manage stress and pain. Breathe in deeply for about 4 counts - feel your stomach expand. Hold that breath for 2 counts and then breath out slowly for 4 counts. Notice the relaxation spread through your body. Try this for a few minutes.”*<br>**Opioid tapering information**<br>*“Opioid medication becomes less effective over time. It might be surprising but after a gradual reduction many people say their pain is more manageable and less distressing.”*<br>*“ Hi [first name], sometimes it takes our body time to adjust to changes in medication. Withdrawal symptoms while reducing your opioid medication are temporary and will pass.”*<br>**Socioemotional support/reassurance**<br>*“It's normal to feel irritable or low while reducing opioids. It's a big change and your body needs time to adjust. Try to be kind to yourself.”*<br>*“[first name], you are doing your best in a challenging situation. Be proud of yourself.”* |

Message delivery will be managed by Twilio (Twilio Inc, San Francisco, California) and will be one-way (i.e., recipients cannot reply directly to messages). Messages will be sent over Australian telephone networks at no cost to individual participants. Messages will be sent at ∼10 AM and ∼4 PM (local time). The messages sent to the participants are standardised in their content and delivery (by day of study and by the time of day; AM/PM). Participants’ first names are used in a selection of messages to increase engagement (e.g., “Hi John…”).

Before watching the educational video and receiving the first daily text messages, participants in the intervention group will receive an information sheet (Appendix 6) outlining the digital support they will receive (i.e., the educational video and daily text messages). Participants will be informed that the text messages are one-way. They can cease receiving text messages by emailing or calling the research team. Participants will be asked whether they are willing to receive daily text messages (i.e., the second consent step). If they do not agree to receive daily text messages, they can continue with just watching the educational video (see *Statistical Methods* for sensitivity analysis).

#### Control group

Participants in the control group similarly will receive a link to an educational video (i.e., active control) to watch after randomisation (day 1) but will not receive daily text messages. The content of the video is identical to the video in the intervention group except that it does not include information/testimonials about receiving daily text messages and the duration is 10 minutes (see *Blinding Arrangements*). Before watching the educational video, participants in the control group will also receive an information sheet (Appendix 6) outlining the digital support they will receive (i.e., the educational video).

### Outcomes

The study primary and secondary outcomes are selected based on the recommendations of the Initiative on Methods, Measurement, and Pain Assessment in Clinical Trials (IMMPACT)^33 34^ To minimise participant burden, we will use short-form versions of previously validated questionnaires as described below and at the times specified in Table 3.

**Table 3:** Data collection plan

| **Table 3: Data collection plan** |  |  |  |  |  |
| --- | --- | --- | --- | --- | --- |
|  | Description | Baseline | Week 4 | Week 8 | Week 12 |
| Demographics –<br>Clinical data | Age, Gender, Socioeconomic,<br>Pain, Medications, Current care,<br>Tapering history | X |  |  |  |
| Decision | Single-item CPSPost | X |  |  |  |
| Social support | OSSS-3 | X |  |  |  |
| Pain | 3-item PEG scale | X | X | X | X |
| Opioid dose, OME | Self-report, prescription history | X | X | X | X |
| Emotional functioning | PHQ-2 and GAD-2 | X | X | X | X |
| Self-efficacy in tapering | Single-item OTSEQ<sup>a</sup> | X | X | X | X |
| Pain cognitions | CAP-6 and PSEQ-2 | X | X | X | X |
| Withdrawal symptoms | Open-ended question |  | X | X | X |
| Current care | Self-report visits for pain | X | X | X | X |
| Global improvement | PGIC scale |  |  |  | X |
| Adverse events | NEQ |  |  |  | X |
| Acceptability | Feedback survey, interview |  |  |  | X |
| Feasibility | Recruitment, dropout, delivery |  |  |  | X |
| Abbreviations: PEG: Pain Enjoyment of life General activity scale; PGIC: Patient Global Impression of Change; CPSPost: Control Preferences Scale; PHQ-2: Patient Health Questionnaire-2; GAD-2: Generalized Anxiety Disorder-2; OME: Oral Morphine Equivalents; CAP-6: Concerns about Pain Scale; PSEQ-2: Pain Self-Efficacy Questionnaire; OSSS-3: Oslo Social Support Scale. <sup>a</sup> OTSEQ will be repeated after watching the video. |  |  |  |  |  |

#### Primary outcome

The primary outcome in this study is the PEG scale score at 12 weeks after enrolment.^29^ The 3-item PEG scale is a subset of the Brief Pain Inventory scale^35^ and has an 11-point NRS for each of the three domains of pain intensity, pain interference with general activity (physical interference), and pain interference with enjoyment of life (affective interference).^36^ Ratings are from 0 (‘no pain’/’does not interfere’) to 10 (‘pain as bad as you can imagine’/’completely interferes’). The total score is the average of the 3 items (ranging from 0 to 10). The minimal clinically important difference (MCID) in the PEG scale has been evaluated in several studies and is reported from 1 to 2.5 points (out of 10). We will consider at least 1 point difference between the two groups in the PEG scale to be a clinically important difference in this study.^29 37 38^

#### Secondary outcome measures

- Emotional functioning 12 weeks after enrolment: We will measure the severity of *anxiety* and *depression* symptoms with the Generalised Anxiety Disorder-2 (GAD-2)^39^ and the Patient Health Questionnaire-2 (PHQ-2),^40^ respectively. Each questionnaire contains 2 items on anxiety/depression symptoms and the responses range from 0 (‘Not at all’) to 3 (‘Nearly every day’). Total score is the sum of the scores for each questionnaire and ranges from 0 to 6. A cut-point score of 3 or greater in each scale is generally used for screening depression/anxiety.^39 40^
- Participant’s rating of global improvement 12 weeks after enrolment: We will measure global improvement using the Patient Global Impression of Change (PGIC) scale, a 7-point Likert scale with ratings ranging from ‘Very much improved’ to ‘Very much worse’.^41^
- Cumulative incidence of withdrawal symptoms 12 weeks after enrolment: We will evaluate the experience of withdrawal-like symptoms every 4 weeks during the study period using a binary question (Yes/No) followed by an open-ended prompt for patients to explain the experienced symptoms.^42^
- Percentage of opioid dose reduction 12 weeks after enrolment: To measure opioid dose reduction, we will use two sources of information: 1) participants’ self-reports of their current medication (i.e., drug names, doses, regimen) at baseline and then every 4 weeks; 2) prescription history obtained from The National Real Time Prescription Monitoring system (RTPM). The RTPM is a nationally implemented system in Australia that collects prescribing and dispensing information about monitored medicines via electronic prescription exchange services. The National Data Exchange component of the RTPM captures information from state and territory regulatory systems, prescribing and dispensing software, and a range of external data sources.^32^ The total daily dose (TDD) of opioids will be converted to mg of oral morphine equivalents (OME) and the percentage of reduction from baseline will be calculated.^43^
- Self-efficacy 12 weeks after enrolment: We will measure pain self-efficacy using the 2-item Pain Self-Efficacy Questionnaire (PSEQ-2) which assesses confidence in the ability to do tasks and activities despite pain with ratings ranging from 0 (‘not at all confident’) to 6 (‘completely confident’) and the total sum score ranging from 0 to 12.^44^ In addition, we will measure self-efficacy in reducing opioid medications using the Opioid Tapering Self-Efficacy Questionnaire (OTSEQ) which we developed based on Bandura’s self-efficacy theory.^45^ The OTSEQ measures self-efficacy to taper prescription opioids in the presence of chronic pain using one item (general self-efficacy). Participants rate their confidence by selecting a number from 0 (‘not at all confident’) to 100 (‘completely confident’). The face validity of the questionnaire has been evaluated by a panel of clinicians during the co-design process and cognitive debriefing was conducted by interviewing six patients with CNCP who had experienced opioid tapering.
- Concerns about pain 12 weeks after enrolment: Pain catastrophising cognitions will be measured using the 2-item Concerns About Pain scale (CAP-6).^46^ Ratings range from 1 (‘never’) to 5 (‘always’) and the total sum score ranges from 6 to 30.
- Acceptability of digital support will be measured using a survey at the end of the study period. The survey design was based on previous studies,^47^ including our pilot RCT,^25^ and contains Likert scales and open-ended questions about the usefulness, readability, and acceptability of the digital support. We will also conduct a semi-structured interview at the end of the study period in both groups to gain a deeper understanding of participants’ experiences and views of the content and delivery of the digital support.
- Feasibility of digital support will be evaluated at the end of the study according to the *number* of messages delivered/not delivered which will be recorded automatically by the text message system. Moreover, *participant disposition* will be reported according to the IMMPACT recommendation^33^ including the recruitment process, adherence, and study completion data, the use of concomitant care (e.g., healthcare visits), and any protocol deviations. This information will be extracted from the data collection software (Research Electronic Data Capture, REDCap).^48^
- Potentially adverse and unwanted events 12 weeks after enrolment: Participants will complete the 20-item Negative Effects Questionnaire at the end of the study. This is a tool commonly used to examine potential adverse and unwanted events in psychological treatments.^49^

#### Other study measures

Demographic (age, gender), socioeconomic (education, employment, marital status, language and English proficiency, state, place of usual residence), and clinical data (pain condition and duration, current care, tapering history) will be collected at the beginning of the study using a self-administered questionnaire. To further explore the factors that may be associated with treatment response, patient involvement in the decision to taper opioids will be measured using the single-item Control Preferences Scale (CPS_Post_ version) which evaluates *perceived* role position after consultation with ratings from 1 (‘I made my decision alone’) to 5 (‘My doctor made the decision’).^50^ The Oslo Social Support Scale (OSSS-3) will be used for measuring perceived social support.^51^

### Assignment of interventions

#### Allocation

We will use the REDCap system for managing enrolment, randomisation, and data collection.^48^ Participants will be enrolled in the study by a Research Assistant (RA). After completing the baseline questionnaires, the RA will use REDCap to randomly allocate the participant to a study group based on the randomisation table embedded in REDCap. The randomisation table will be generated at the beginning of the study by an independent member of our Clinical Trial Unit using Research Randomizer (https://HYPERLINK "http://www.randomizer.org/)" www.randomizer.org/). Access to this table in REDCap will be limited for the research team members. The RA can only see the assigned group after (but not before) the allocation (concealed allocation) as this is required for managing the delivery of interventional text messages via the text message system.

#### Blinding/Masking

Participants, their clinicians, data collectors, research team members (except the RA who manages the text messaging system), and statisticians are all blinded to the group allocation. Participants will be informed by the PIS (Appendix 3) that they will be randomly allocated into two groups and that both groups will receive ‘digital health support’. They will be informed that the study’s aim is to evaluate whether digital health support can improve health outcomes in patients with chronic pain during reducing opioid medications. This *limited disclosure* of the study aim is required to maintain blinding (masking) but does not involve deception.^30^ This method is recommended for “Improving Blinding Integrity and Reporting in Psychotherapy Trials”.^52^ After randomisation and before receiving the digital support, participants in both groups will become aware of the details of the digital support that they will receive but will not become aware of the other group (Appendix 6). Participants will be asked not to discuss the details of the digital health support they receive with the clinician supervising their opioid tapering for the duration of the study.^52^ The study RA will not be blinded to the group allocation but will not be involved in data collection after randomisation. Questionnaires will be completed online (self-administered). For other data (e.g., feasibility data) groups are not known to the data collectors.

### Data collection and management

Participants will complete questionnaires developed in REDCap and delivered online at the time points specified in Table 3. The link to the REDCap surveys will be sent to the participant’s email address and text messages will be sent to their mobile phone as a notification and, if needed, as a reminder to complete the surveys.^53^ Semi-structured interviews will be conducted over the phone at the end of the study (week 12). The interviews will be audio-recorded and transcribed. Participants will be invited for interview, consecutively as they finish the study until a diverse range of in-depth experiences are captured.^54^ Feasibility data including the number of messages delivered will be extracted from the SMS software. Data including enrolment, refusals, drop-outs, and withdrawal from the study will be extracted from REDCap records.

### Statistical methods

#### Sample size

With a 2-sided α level of 0.05, a study power of 0.8, a standard deviation (SD) of 2.5 points in the PEG scale score (primary outcome measure), and assuming a 10% drop-out rate, we require 220 participants in total to detect 1 point difference (MCID of PEG, Cohen’s *d* = 0.4) between the two study groups in mean PEG score after 12 weeks.

#### Statistical analysis

Researchers conducting analyses will be blinded to the group status. Our primary outcome is the PEG scale total score at 12 weeks post-randomisation. The study primary hypothesis (H1) is that the PEG scale total score at 12 weeks after randomisation is lower in the intervention compared to the control group. To test this hypothesis, a linear mixed model containing fixed effects of treatment group, time, and time x group interaction as well as baseline values of the outcome, will be used. A planned contrast (i.e., the between-group difference at week 12) will be used to test the study primary hypothesis. We will also include the measures of the PEG scale at 4 and 8 weeks after randomisation in our primary analysis model, so that in the case of missingness at 12 weeks, the model can use these data to obtain a better estimate despite the missingness by applying maximum likelihood methods (i.e., including the random effect of participants and fixed effect of time). This approach provides valid inferences under the assumption that the mechanism of missing data is ignorable (or Missing At Random). To account for the correlation between repeated measures at 4, 8, and 12 weeks, a random intercept will be included at the subject level. We will explore if the baseline adjusted model (i.e., including baseline values of the dependent variable in the model as a covariate) can improve the precision of the estimates based on the fit statistics and residual distribution. Normal distribution of the residuals will be tested. If departing from normality, first we will look for a more appropriate distribution shape and data will be transformed if appropriate to approximate residuals to normality.

Similarly, a linear mixed model will be used to analyse the study secondary outcomes with repeated measures after randomisation (e.g., emotional functioning). To analyse continuous clinical secondary outcomes, we will use the same model as the primary analysis. This involves linear mixed models that incorporate outcome measures at all available time points, along with a group x time interaction term that enables estimation of the difference between arms at each time point. For binary secondary outcomes, we will employ a logistic mixed effects model, again including the outcome at all available time points and with two levels. The statistical significance will be defined as *P* < 0.05 (two-sided) and the effect estimates will be reported with corresponding Confidence Intervals (CIs). Cohen’s *d* (effect size) will be calculated based on the estimates and standard errors. Analyses will be conducted using SAS software (v 9.4, SAS Institute Inc., Cary, NC, USA).

#### Explorative analysis

The PEG scale total score is the primary outcome measure to test the efficacy of the intervention according to the pre-specified MCID for this measure. However, each of the PEG scale items will also be analysed and reported separately with explorative aims. Explorative analyses will be done to investigate trends of change in outcomes within- and between-group (i.e., group x time interaction effect) and potential moderating and mediating factors.^55^ Model statement and parameters (i.e., estimation method, covariance structure) will be determined based on the fit statistics and residuals’ distribution. In addition to marginal linear mixed models (where time is a categorical variable), linear mixed models (where time is a continuous variable) with random intercept and random slope will also be conducted. Pairwise contrasts will be done to compare the outcomes between the two groups at weeks 4 and 8. In these explorative analyses, *P* value adjustment will be done to account for multiple comparisons according to the Holm–Bonferroni method.^56^

#### Sensitivity analyses and missing data imputation

Sensitivity analyses will be done with both per-protocol and intention-to-treat (ITT) analyses based on drop-outs and missing data, as well as based on participants’ compliance (e.g., refusing to receive SMS if allocated to).^57^ For the analyses where all levels of the repeated measures are included in the model as the dependent variable, missing data will be imputed by the mixed-model analysis without any further ad hoc imputation unless when only a baseline value is available.^58^ In such a case, selective imputation will be done using the baseline observation carried forward method.^57^

#### Qualitative analysis

For feasibility and acceptability outcomes, a mixed-method analysis will be conducted on quantitative-qualitative data. Transcripts and digital notes will be entered into NVivo (version 12, QSR International). Thematic analysis using an inductive approach will be followed. The Consolidated Framework for Implementation Research will be used to organise implementation barriers and facilitators into major constructs.^59^

## Ethics and dissemination

### Research ethics approval and protocol

This study has approval from the Northern Sydney Local Health District Research Ethics and Governance (2022/ETH01795, 2022/STE02938). The initially approved protocol version was 1.02 (dated 11 October 2022). The protocol has been revised before enrolment (version 1.3 dated 19 January 2023). The trial was prospectively registered at the Australian New Zealand Clinical Trials Registry (ACTRN: ACTRN12622001423707).

### Dissemination

Study results will be published in peer-reviewed journals and presented at scientific and professional meetings.

## Discussion

This study describes the protocol for a double-blind RCT to test the efficacy of a co-designed digital health intervention to support people with CNCP during tapering LTOT. Other outcomes include feasibility, acceptability, and safety of the intervention. The intervention content and the trial protocol have been co-designed in collaboration with patients with chronic pain and clinicians specialising in pain management. If proven to be efficacious, this digital support can be a low-cost intervention to be implemented at a large scale.

There are limitations in the design of this trial that should be acknowledged. We acknowledge that there may be substantial variation in opioid dose changes attempted by participants over the 12-week study period, and there is likely to be significant variation in participants’ satisfaction, engagement with, and adherence to the ‘usual care’. However, heterogeneity in participants’ opioid tapering schedules and usual care will allow us to capture potential feasibility challenges. Moreover, it would be impractical and potentially not beneficial to the principle of patient-centred care to standardise ‘usual care’ or standardise opioid tapering schedules.^10^ We also acknowledge that our calculation for the percentage of opioid dose reduction is based on self-report of opioid consumption, which is prone to bias. This method of assessment reflects the reality of clinical care delivered in the current study setting (Australia), where patients’ adherence to opioid prescribing and tapering advice is not enforced or closely supervised but relies on patient self-report. To minimise potential bias, we will collect prescribed/dispensed opioid medication data from electronic monitoring systems as a second source of information. Also, to minimise the risk of social desirability effects on opioid dose reporting, participants are reminded that their assessments, including medication reporting, will be confidential and not reported back to their clinician. We also acknowledge the study limitation regarding the control condition. Designing a sham or placebo digital health is challenging for this study. Therefore, we used an active control condition (an educational video) and the *limited disclosure* method to achieve masking. However, the educational video is likely to provide benefits to the participants as it has been shown to improve proximal outcomes including perceived tapering effectiveness and tapering self-efficacy,^21^ and can promote health behaviour change.^22^ This may lead to an underestimation of the actual effect size of the digital support.

## Data Availability

NA, this is a study protocol

## Acknowledgements

We acknowledge the Ernest Heine Family Foundation for supporting this study with a philanthropic gift to the University of Sydney. MLF holds an NHMRC Investigator Grant.

## Authors Contributions

PG and CEAJ conceptualized the interventions and acquired funding and should be considered joint senior author. All authors contributed to the design of the study protocol. AG drafted the study protocol and manuscript. All authors critically reviewed and edited the manuscript and approved its final version.

## Supplementary Materials

Appendix 1 - The SPIRIT checklist.

Appendix 2 - Study Advertisement Materials.

Appendix 3 - Participant Information Statement and Consent Form.

Appendix 4 - Screening form.

Appendix 5 - Text messages and Video script.

Appendix 6 - Instruction For Digital Health Support.

