## Appendix 2 for "A text messaging intervention to support patients with chronic pain during prescription opioid tapering: protocol for a double-blind randomised controlled trial"

**Study Advertisement Materials**

Study Flyer

This research has been approved by the NSLHD HREC reference number 2022/ETH01795

### Is chronic pain impacting your everyday life?

**If you are currently using opioids for pain, this study may be for you**

#### Digital health support for adults with chronic pain reducing opioids

Researchers from The University of Sydney are looking for adults with chronic pain to participate in an innovative study using digital health to support people to reduce their opioid medication dose.

#### What does it involve?

- Using one of the digital health interventions
- Completing monthly surveys for 3 months
- An interview about your experiences with the intervention

#### Chief Investigator:

Prof. Paul Glare

#### Contact us


Phone: [dedicated phone number]

#### Location

This study is conducted online and over phone. You can participate from anywhere in Australia provided you have access to a phone network and the internet.

#### Are you eligible?

- 18 years or older
- Living with chronic pain
- Using opioids and planning to reduce your dose (or already reducing)
- Willing to provide your doctor's contact details

**Scan this QR Code to find out more**

**XXX**

[link to study webpage]

Twitter & Facebook post

Support4pain is an innovative study using Digital Health to support people with chronic pain while they are reducing their opioid dose. You can participate from anywhere in Australia provided you have access to a phone network and the internet. Follow http://XXXX to find out more! This research has been approved by the NSLHD HREC reference number 2022/ETH01795

**Do you have CHRONIC PAIN?**
**Are you about to reduce your OPIOID DOSE?**

Researchers from The University of Sydney are looking for people with **CHRONIC PAIN** to test whether **DIGITAL HEALTH** can support them while they are **REDUCING THEIR OPIOID DOSE**.

QR Code

XXX

[link to study webpage]

Follow the link

XXX [link to study webpage]

Study webpage

### Digital Health Support for Opioid Dose Reduction

Researchers from The University of Sydney are looking for adults with chronic pain to participate in this innovative study which will use digital health to support opioid dose reduction.

#### Study description/purpose

Many people with chronic pain use opioid medications long-term. Research shows the harms associated with these medications can outweigh the benefits when used for chronic pain. Reducing opioid medications not only minimises the harms but can also lead to improvements in pain and quality of life.

Researchers at The University of Sydney are conducting an innovative study on **DIGITAL HEALTH** to support people with **CHRONIC PAIN** reducing their **OPIOID** dose. They are looking for adults with chronic pain who have started or are going to start to reduce their opioid dose under the supervision of their doctor. Participants can join this study from anywhere in Australia where they have access to a phone network and the internet.

#### What is involved?

Participants will continue their care as usual with their doctor and other healthcare professionals and will reduce their dose under their supervision. In addition, they will receive one of the digital health interventions provided by this study and complete surveys every month for 3 months. You can read more about this study in the Participant Information *here* [link to Participant Information Sheet]

**Am I eligible?** Please fill out a short survey *here* [link to screening form] to find out

**Chief investigator:** Prof. Paul Glare

**Institutions:** The University of Sydney, Pain Management Research Institute. This research has been approved by the NSLHD HREC reference number 2022/ETH01795.
