## Appendix 3 for "A text messaging intervention to support patients with chronic pain during prescription opioid tapering: protocol for a double-blind randomised controlled trial"

### PARTICIPANT INFORMATION SHEET AND CONSENT FORM

#### Digital Support For People With Chronic Pain Who Are Reducing Prescription Opioids

| | |
|---|---|
| **Project Sponsor:** | University of Sydney |
| **Principle Investigator:** | Professor Paul Glare |
| **Associate Investigators:** | Associate Professor Claire Ashton-James |
| | Dr Ali Gholamrezaei |
| | Michael Magee |
| | Amy Gray McNeilage |

**Location:** Pain Management Research Institute, Ground floor, Douglas Building, Royal North Shore Hospital, St. Leonards NSW 2065

##### 1. Invitation

We invite you to participate in research about digital support for patients with chronic pain who are reducing their opioid dose. Before you decide whether you wish to participate in this study, it is important for you to understand why the research is being done and what it will involve. Please take the time to read the following information carefully and discuss it with others if you wish.

##### 2. 'What is the purpose of this study?'

This research evaluates whether providing education, motivation, and emotional support using digital health technologies can benefit patients with chronic pain who are reducing their opioid medications and improve their pain and well-being.

##### 3. 'Why have I been invited to participate in this study?'

You are invited to participate in this study if you are currently prescribed opioid medications for chronic pain, and either your doctor has recommended that you reduce your opioid dose, or you are currently reducing your opioid dose under the supervision of your doctor.

##### 4. 'What if I don't want to take part in this study, or if I want to withdraw later?'

Participation in this study is voluntary. It is completely up to you whether you participate. If you decide not to participate, it will not affect your relationships with your treating clinicians or the treatment you receive now or in the future.

If you wish to withdraw from the study, you can do so at any time without having to give a reason.

If you choose to withdraw, you can inform the research team by sending an email to or by phone call to [dedicated phone number] during office hours. They will send you the '**Participant Withdrawal of Consent Form**' to sign.

If you withdraw from the study, your data and information will be deleted from study records and will not be used in future research, unless you agree to your de-identified (anonymised) data being used.

Note: The principal investigator may remove a participant from the study at any time if they feel it is in their best interest or that their safety may be at risk due to unanticipated health issues or not following study procedures.

#### 5. 'What does this study involve?'

Your participation in the study will last for 12 weeks starting from the day you sign the consent form.

If you decide that you would like to participate, the following will happen:

1. You will complete a short online eligibility form. This will take around 5 minutes to complete. If you are eligible to participate, you will be asked to provide your contact information. A member of our research team will then contact you by email to confirm your eligibility.

2. If you are eligible to participate, you will be asked to provide the contact details of your doctor (GP or Pain Specialist) who is supervising your opioid medication reduction. The research team will contact your doctor to inform them that you are participating in this research. This is a requirement of the study. However, data collected during the study will not be reported to your doctor. A copy of the letter your doctor will receive is available at this link [link to PDF file of Appendix 5_GP_Specialist Letter].

3. The research team will send you the consent form to sign online. As part of the consent, we will ask for your permission to access your prescription history in the National Real Time Prescription Monitoring system. This is a national database that gives prescribers and pharmacists real-time information about your prescribing and dispensing history for certain medicines such as opioids. This will help them to make safer decisions about your care. To find out more about this (including privacy and data protection considerations] you can visit this website [link to https://www.health.gov.au/our-work/national-real-time-prescription-monitoring-rtpm).

   Please note, this is a requirement for participating in this study to give us permission to access your prescription history so that we can verify that opioid medications are prescribed for you. We will record your prescribed opioid medications once at the beginning of the study (only the last 4 weeks prescriptions) and once at the end of the study (only the last 12 weeks prescriptions). This will help us to evaluate whether the digital health support is helpful in reducing the prescribed opioid medications.

   A research team member who is a qualified medical practitioner will use SafeScript NSW [link to https://www.safescript.health.nsw.gov.au/consumers] to access your prescription history. No other information will be obtained from this database.

4. Once you have signed the consent form, the research team will send you an online survey with questions such as about your age, gender, pain, and health status. The baseline survey will take around 15 minutes to complete.

5. As part of this study, you will be randomly assigned to receive one of two digital health supports. This is done by a computer but is similar to flipping a coin. The digital health supports, delivered by mobile phone and email, are designed by a team of researchers, patients, and clinicians. They provide education, motivation, and emotional support to people with chronic pain while reducing their opioid medications. You will receive more details about the digital support once it is known which you will receive.

6. This is a double-blind study. This means that researchers, participants, and participants' doctors will not know which digital support the participant is receiving. You will be asked not to talk to your doctor or the researchers you are in contact with about the details of the digital support you are receiving unless it is directly requested by your doctor or a research team member or you decide to do so for safety reasons.

7. This digital health support is not replacing any care you would otherwise receive from your doctor or other healthcare professionals. It is provided in addition to your usual care, and you will continue your treatment under their supervision.

8. You will complete short questionnaires every month for 3 months that ask about your pain and other symptoms, function, mood, thoughts, and medication changes. Each monthly survey will take around 5 minutes to complete. These help us to evaluate whether the digital support is helpful. These will be sent to your mobile phone and/or email and will be completed online. You will receive text messages as reminders to complete the questionnaires.

9. After 12 weeks, you will be asked to complete a feedback survey and may be invited to participate in an interview over the phone (audio-recorded) which will take around 30 minutes to give feedback about the digital support you received. Participating in the feedback interview is optional and not a requirement of the study.

**Please note, that the digital support in this study is not a crisis service and if you are in crisis or it is a health emergency, you should contact your nearest hospital or an emergency health service available in your area. If you experience a flare-up of pain or have any other pain-related concerns, you should contact your doctor or clinician as you would normally.**

#### 6. 'How is this study being paid for?'

This study is supported by a grant to The University of Sydney from the Ernest Heine Family Foundation.

#### 7. 'What are the alternatives to participating in this study?'

The digital health support is not replacing the care that you are receiving from your doctor and other healthcare professionals. If you decide not to participate, your treatment will continue as usual.

8. ‘Are there risks to me in taking part in this study?’

We do not expect you to experience any harm as a result of participating in this study. If any unexpected consequences occur during your participation or you feel distressed by this study, please contact the research team by email address for advice, or contact your GP, or attend your local Hospital Emergency Department.

We also encourage you to reach out to the following services if needed:

- For mental health support, call Lifeline: 13 11 14 (24 hours)
- For withdrawal support, contact the Alcohol and Drug Information Service: 1300 340 357 (24 hours)
- For chronic pain support, call the Pain Link Peer Support Service: 1300 340 357 (leave a message for call back service)

**Please note, that the study phone number [study phone number used by the SMS software] cannot be contacted by call or text messages. The email address and other study phone number [dedicated phone number] are NOT for urgent medical/mental health needs.**

9. ‘What happens if I suffer injury or complications as a result of the study?’

If you suffer any injuries or complications because of this study, you should contact the research team by the provided email address. They will assist you in arranging appropriate medical treatment as soon as possible.

You may have a right to take legal action to obtain compensation for injuries or complications resulting from this study. Compensation may be available if your injury or complication is caused by the interventions in this study, or by the negligence of any of the parties involved in the study. If you receive compensation that includes an amount for medical expenses, you will be required to pay for your medical treatment from those compensation payments.

If you are not eligible for compensation for your injury or complication under the law but are eligible for Medicare, you can receive medical treatment free of charge in any Australian public hospital.

10. ‘Will I benefit from the study?’

The digital health supports in this study have previously been shown to provide benefits to patients with chronic pain reducing their opioid medications. Therefore, you may see a reduction in your symptoms and/or improvement in function and/or mood. However, we cannot guarantee this, and you may not benefit from the study. This research will also help us to better understand how digital health supports can help patients with chronic pain reduce their opioid medications and to develop programs that may improve access to treatment for people with this condition.

11. ‘Will taking part in this study cost me anything, and will I be paid?

The digital health support will be provided to you for free. However, your participation in this study will cost you some time and energy including the time it takes to complete the study surveys and attending in an interview over the phone which, in total, will be around 75 minutes throughout the study. You will not receive payment to participate in this study.

#### 12. 'How will my confidentiality be protected?'

Identifiable information (name and contact details) will be collected only if you are eligible to participate in the study. At the beginning of the study, the research team will contact your doctor to inform them that you are participating in this research. However, data collected during the study will not be reported to your doctor. Any identifiable information that is collected about you in connection with this study will remain confidential and will be disclosed only with your permission, or except as required by law. Only the researchers named above and, if required for monitoring purposes, The Northern Sydney Local Health District Human Research Ethics Committee and regulatory bodies will have access to your information and data, which will be held securely at The Pain Management Research Institute.

We will contact you from an email address specifically used for this study. We will send text messages to your phone using this number [study phone number used by the SMS software] and over Australian telephone networks at no cost. We will use REDCap software for online surveys which will be saved on The University of Sydney server. These are automated and secure systems with protocols for encryption and authentication.

Recorded interviews will be transcribed by Rev transcription services. Rev professionals have all signed Non-Disclosure Agreements and strict confidentiality agreements, and complete transcription work on a secure platform that encrypts client files using protocols at bank-level security. The Rev Privacy Policy can be found at https://www.rev.com/about/privacy.

#### 13. 'What happens with the results?'

Data collected online will be automatically saved in a database on the University of Sydney protected server. Paper documents (if any) will be scanned and uploaded into this database. Paper documents will be stored at the Pain Management Research Institute in locked cupboards. Audio files from interviews and transcripts will be saved in a password-protected computer owned by the University of Sydney to ensure data security. Data within the SMS software will be deleted after being exported at the study closure. Exported data from the database will be de-identified and stored in a password-protected computer owned by the University of Sydney to ensure data security and confidentiality. Data gathered in this study may be used by the research group of this study in future research approved by an ethics committee and such data will not contain your name or any identifiable information about you. All data will be stored for 15 years after the publication of the project's final report and then will be securely destroyed per requirements for clinical research.

We hope that the results of this research project will be published in scientific journals and presented at conferences and other meetings. All data that is collected in this study, including data collected by the online eligibility form from all applicants, will be saved and de-identified to remove personal information before analysis and reporting. In any publication, report, or presentation, information will be provided in such a way that you cannot be identified, except with your permission. The overall results of the study will be provided to you if you wish.

##### 14. 'What happens to my treatment when the study is finished?'

After the study is finished, your care will continue as usual. You can save and access the information sent to your phone and email for as long as you wish.

##### 15. 'What should I do if I want to discuss this study further before I decide?'

##### 16. 'Who should I contact if I have concerns about the conduct of this study?'

This study has been approved by the Northern Sydney Local Health District HREC. Any person with concerns or complaints about the conduct of this study should contact 02 9926 4590 or and quote HREC reference [2022/ETH01795 and 2022/STE02938].

**Thank you for taking the time to consider this study.**

**If you wish to take part, please read and sign the consent form provided on the next page.**

**You can keep a copy of this information sheet.**

Pain Management Research Institute

Ground floor, Douglas Building, Royal North Shore Hospital, St. Leonards NSW 2065

### CONSENT FORM

#### Digital Support For People With Chronic Pain Who Are Reducing Prescription Opioids

1. I agree to participate as a participant in the study described in the Participant Information Sheet set out above.

| |
|---|
| Please provide your full name |
| Please provide your date of birth |

Please ensure the full name and date of birth are correct.

2. I have read the Participant Information Sheet explaining why I have been selected, the aims of the study, and the possible risks, and the study has been explained to me to my satisfaction.

◯ YES / ◯ NO

3. I confirm that reducing my opioid dose will be under the supervision of my doctor (my GP, Family Doctor, or other Specialist Physicians).

◯ YES / ◯ NO

4. I agree that the research team contact my doctor and inform them about my participation in this study.

◯ YES / ◯ NO

5. As it is explained in the participant information sheet (page 2), it is a requirement of participating in this study that we access your prescription history in the National Real Time Prescription Monitoring database so that we can verify you are prescribed opioid medications and to record your prescribed opioid medications once at the beginning of the study (the last 4 weeks prescriptions) and once at the end of the study (the last 12 weeks prescriptions). This will be done by a research team member who is a qualified medical practitioner using SafeScript NSW. No other information will be obtained from this database.

I agree that a research team member who is a qualified medical practitioner to access my prescription history in the Real Time Prescription Monitoring database as specified above.

◯ YES / ◯ NO

6. Before signing this consent form, I have been given the opportunity to ask any questions relating to possible physical and mental harm I might suffer as a result of my participation, and I have received satisfactory answers.

◯ YES / ◯ NO

7. I understand that I can withdraw from the study at any time without affecting my relationships with the research team or my treating clinicians.

◯ YES / ◯ NO

8. I agree that research data gathered from the study may be published and that publications will not contain my name or any identifiable information about me.

◯ YES / ◯ NO

9. I agree that data gathered in this study may be used by the research group of this study in future research approved by an ethics committee and that such data will not contain my name or any identifiable information about me.

◯ YES / ◯ NO

10. I understand that if I have any questions relating to my participation in this research, I may contact the research team by email at or by phone [dedicated phone number] during office hours who will be happy to answer them.

◯ YES / ◯ NO

11. I acknowledge receipt of a copy of this Consent Form and the Participant Information Sheet.

◯ YES / ◯ NO

**'I understand that by submitting this consent form I consent to participate in the study as outlined in the Participant Information Statement.'**

◯ YES / ◯ NO

**SUBMIT**

**Please note, that if you have a question about your pain management or medications, you need to contact your GP or clinician as you would normally.**

If you have a question relating to the conduct of this study such as information provided in the Patient Information Sheet and Consent form or completing this consent form or to withdraw from the study at any time, please contact the research team by or by phone [dedicated phone number] during office hours.

Please indicate below if you would like to be informed about the overall results of this study when available. This will be sent to your email address.

I would like to be informed about the overall results of this study when available.

◯ YES / ◯ NO

Pain Management Research Institute

Ground floor, Douglas Building, Royal North Shore Hospital, St. Leonards NSW 2065

#### REVOCATION OF CONSENT

### Digital Support For People With Chronic Pain Who Are Reducing Prescription Opioids

I hereby wish to **WITHDRAW** my consent to participate in the study described above and understand that such withdrawal **WILL NOT** jeopardise any treatment or my relationships with my doctors or my medical attendants or with the researchers.

For online form

Please provide your full name

**'I understand that by submitting this withdrawal form I am withdrawing my consent to participate in the study as stated in the information statement.'**

◯ YES / ◯ NO

**'I agree to my de-identified (anonymised) information and data being used in this study and/or future research.'**

◯ YES / ◯ NO

**SUBMIT**

For paper form

Signature: Date: / /

The section for Revocation of Consent should be forwarded to:

**Chief Investigator**

**Professor Paul Glare**

Pain Management Research Institute, Ground floor, Douglas Building, Royal North Shore Hospital, St. Leonards NSW 2065
