## Appendix 4 for "A text messaging intervention to support patients with chronic pain during prescription opioid tapering: protocol for a double-blind randomised controlled trial"

**Screening form**

**Digital Support For People With Chronic Pain Who Are Reducing Prescription Opioids**

[Please note that the information in *[]* are referring to how this form is designed in the REDCap and branching logic function, and are hidden to the participants]

Thank you for your interest in this study. To determine whether this study is right for you, please answer a few questions below. All information collected in this study will remain confidential. Please ensure you have read the Participant Information Sheet here [Link to Participant Information Sheet].

**<u>Please Note:</u>** Participating in this research may not be suitable for people with any of the following conditions:

- *Opioid or other substance use disorder*
- *Bipolar disorder, psychotic disorder, schizophrenia, or personality disorder*
- *Suicidal thoughts or behaviour*
- *Those who have taken illicit drugs or someone else's prescription pain medication in the past 12 months*

We always strongly recommend people discuss their participation in this research with their GP (family doctor) and other treating doctors.

◯ **I confirm that none of the above conditions applies to me.**

[Proceed with the next steps only if the participant has selected the confirm button]

[Step 1]

1. Do you currently have pain that has been present for more than 3 months?
   - Yes / No

2. Have you been using prescribed opioids for your pain for one month or more? (example.g., codeine, oxycodone, tramadol, tapentadol, morphine, buprenorphine, methadone, fentanyl)
   - Yes / No

3. Has your doctor (GP or pain specialist) advised you to reduce your opioid dose?
   - Yes / No

4. Are you planning to reduce your opioid dose under the supervision of your doctor (GP or pain specialist)?
   - Yes / No

5. Will you be living in Australia for the next 3 months?
   - Yes / No

6. Are you 18 years or older?
   - Yes / No

7. Are you currently pregnant or planning to become pregnant in the next 3 months?

- Yes / No

8. Have you had any major surgery in the past 6 months or plan to do in the next 3 months (e.g., transplant, removal of a tumour, kidney surgery, heart surgery, surgery on the spine or knee)?
- Yes / No

9. Are you able to read and understand the English language well?
- Yes / No

10. Are you currently participating in another clinical trial?
- Yes / No

11. Are you willing to provide us with the contact details of your doctor (GP or pain specialist)? We need to inform them about your participation in this study.
- Yes / No

[If the criteria in Step 1 are NOT met, present Message #1. Participant cannot be included in the study]

[Message #1]

- Thank you for completing this form. Based on your responses, participating in this research may not be suitable for you. Unfortunately, we cannot include you in this study.
- We encourage you to seek help wherever you feel most comfortable. This might be your GP, family or friend, religious or community leader, or anyone you feel you can trust.
- If you are searching for local health and community services support but don't know where to start, see this Service Finder [https://www.lifeline.org.au/get-help/service-finder/].
- For chronic pain support, call the Pain Link Peer Support Service: 1300 340 357 (leave a message for call back service).
- For mental health support, call Lifeline by 13 11 14 (available 24 hours) or visit the Lifeline website [https://www.lifeline.org.au/]
- For withdrawal support, contact the Alcohol and Drug Information Service: 1300 340 357 (available 24 hours)

[Step 2: If the criteria in Step 1 are met, proceed with screening for a major psychiatric disorder and suicide risk using the following questions]

1. Do you have a poorly controlled or unstable mental illness?
Yes / No

2. Have you planned for or attempted suicide in the past 12 months?
Yes / No

3. Are you at risk of suicide?

Yes / No

4. Have you taken illicit drugs or someone else's prescription pain medication in the past 12 months?
Yes / No

[If reply ‘Yes’ to question 1 or 4, but ‘No’ to question 2 and ‘No’ to question 3, present Message #1. If reply ‘Yes’ to question 2 and ‘No’ to question 3, present Message #2. Participant cannot be included in the study in any of these cases]

[Message #2]

- Thank you for completing this form. Based on your responses, participating in this research may not be suitable for you. Unfortunately, we cannot include you in this study.
- We encourage you to seek help wherever you feel most comfortable. This might be your GP, family or friend, religious or community leader, or anyone you feel you can trust.
- If you are searching for local health and community services support but don't know where to start, see this Service Finder [https://www.lifeline.org.au/get-help/service-finder/].
- We encourage you to call Lifeline by 13 11 14 (available 24 hours) for mental health support or visit the Lifeline website [https://www.lifeline.org.au/].
- For chronic pain support, call the Pain Link Peer Support Service: 1300 340 357 (leave a message for call back service)
- For withdrawal support, contact the Alcohol and Drug Information Service: 1300 340 357 (available 24 hours)

[If reply ‘Yes’ to question 3, ask questions 3b]
3b. Over the last 7 days have you had any suicidal thoughts?
Yes / No

[If NO -> present Message #2, If YES -> proceed with question 3b]

3c. Do you have any current intention or plan of harming yourself?

[If NO -> present Message #2, If YES present Message #3]

[Message #3]
- Thank you for completing this form. Based on your responses, participating in this research may not be suitable for you. Unfortunately, we cannot include you in this study.
- We encourage you to urgently see your GP if that is possible for you or go to your local hospital for mental health support. If none of these are possible for you, please urgently call Lifeline 13 11 14 (available 24 hours) or Suicide Call Back Service 1300 659 467 (available 24 hours). If life is in danger, call 000.
- If you are searching for local health and community services support but don't know where to start, see this Service Finder [https://www.lifeline.org.au/get-help/service-finder/].

[Step 3: If eligible in all steps, present Message #4]

[Message #4]
Thank you for completing this form. Based on your above responses, you may be eligible to participate in our study.

1. Please provide us with your contact details below and you will be contacted by a member of our research team:
Name: …
Email: …
Mobile phone number: …

2. Please provide us with the contact details of your doctor (GP or pain specialist):
Name: …
Email: …
Contact number: …

3. How did you find out about this research?
- o Social media (Facebook or Twitter)
- o Newspaper
- o Printed flyer in a clinic or hospital
- o Email
- o Family/friends
- o My doctor or clinic staff
- o Other, please explain

We appreciate and value your time and effort. We aim to process all applications within 3 business days. However, sometimes we receive a lot of applications, and can be delayed - we will let you know if we expect to be delayed.

This study has been approved by the Northern Sydney Local Health District HREC. Any person with concerns or complaints about the conduct of this study should contact 02 9926 4590 or and quote HREC reference [2022/ETH01795].
