## Appendix 5 for "A text messaging intervention to support patients with chronic pain during prescription opioid tapering: protocol for a double-blind randomised controlled trial"

**TEXT MESSAGES (SELECTED)**

1. a Welcome to our study! We will send you a link to a short video for you to watch later today. It is important that you watch this video as it will provide you support for pain management.  We will send you two SMS per day throughout the study providing you with continued support and reminders to complete online surveys.

1. b Welcome to our study! We will send you a link to a short video for you to watch later today. It is important that you watch this video as it will provide you support for pain management. We will send you SMS reminders throughout the study to complete online surveys.

2. a Hi [first name]. We have sent a link to a short video to your email. The video includes information on pain management that you might find helpful while reducing your opioid dose. It also includes some important information about the study. Please watch the video as soon as you are able.

2. b Hi [first name]. We have sent a link to a short video to your email. The video includes information on pain management that you might find helpful while reducing your opioid dose. It also includes some important information about the study. Please watch the video as soon as you are able.

3. Hi [first name], There are many ways to manage pain without opioids and there are lots of resources to help with pain self-management online. Here's a good place to start: https://aci.health.nsw.gov.au/chronic-pain.

4. Hi again [first name]. When people reduce their opioids, they are able to think more clearly and do more to look after themselves and others.

5. Pain flare ups are like bad moods. Sometimes they come on for no clear reason. They will often pass if you distract yourself. Try doing or thinking about something you enjoy.

6. Withdrawal symptoms can be reduced when you look after your health. Good nutrition, sleep, and exercise will help. It also helps to reduce your dose very slowly.

7. … 168

169. a Hi [first name]. Thank you again for participating in the study. We have sent a feedback survey to your email. Your feedback will help us to improve the way we support people with chronic pain who are reducing their dose of opioids.

THE UNIVERSITY OF SYDNEY

Health
Northern Sydney
Local Health District

**VIDEO SCRIPT (SELECTED)**

You are watching this video today because your doctor has recommended you reduce your opioid medication dose. This can be a daunting prospect and many people find that it helps to have information and support. This video presents a few strategies for managing your pain and maintaining your health and well-being as you reduce your dose of prescription opioid medications. What helps? Understanding the meaning of your pain helps. Knowing that chronic pain is more about sensitivity than damage can help reduce the worry that often goes hand in hand with the experience of chronic pain. Chronic pain is a bit like an over-responsive smoke alarm that goes off when you are cooking or having a steaming hot shower. The loudness of the alarm is real and distressing but there's no fire or real danger that we need to protect ourselves from…

[P1] I've felt so much better to be off the tablets, absolutely wonderful. I've felt stronger, more life you know, more energy. It didn't hurt nowhere near as much so I've felt absolutely great and after that I've learned to deal with the pain itself without the painkillers. I didn't feel I had a life, I felt very enclosed, low, constantly tired, just I didn't want to socialize because I was so tired and rundown and in pain. Once you know what a flare-up is and how to deal with it, it becomes a lot less scary and severe and you just go 'oh yeah I'm having a flare up, all right, let's deal with this'.

[P2] … [P4]

[only group 1] In addition to this, we will support you by sending you a couple of text messages a day over the next four weeks to provide you with information and strategies that have been helpful for others. Each text message has been written together with patients who have experience reducing opioid pain medications and they have been also endorsed by clinical experts. Researchers have found that receiving daily text messages can really help people to change their behaviour and habits. Here's a few individuals telling us about their experiences using the digital support.

[P5] The messages were wonderful… they helped me immensely. They just kept pushing me along… It made me feel that it was okay that I felt like that… They gave me support when I needed it and I could keep reading them all day.

[P6] …

We look forward to hearing about your experiences.
