## Appendix 6 for "A text messaging intervention to support patients with chronic pain during prescription opioid tapering: protocol for a double-blind randomised controlled trial"

THE UNIVERSITY OF SYDNEY

Health
Northern Sydney
Local Health District
NSW GOVERNMENT

### INSTRUCTIONS FOR DIGITAL HEALTH SUPPORT

Thank you for participating in our study, Digital Support For People With Chronic Pain During Prescription Opioid Tapering. The information provided here explains how this digital health support works.

**This digital health support is provided to you <u>in addition</u> to the care and treatments that you are receiving from your doctors and other healthcare professionals. Please continue to follow their advice.**

You will receive a text message (SMS) from this phone number [study phone number]. Please save this number in your contact list using the name **Support@USYD**.

The text message has a link to a video. You will be asked to watch this video. We will also send it to your email address. You can watch it on a computer or tablet if you like. Check your email for the link once you have received the text message. The link to the video will remain active. You can watch it again if you like.

The duration of the video is about 10 minutes. The video has information about:

- Chronic pain and opioid medications
- What to expect while reducing opioid dose?
- How reducing opioid dose can help you?
- Pain management techniques that can help you while reducing opioid dose
- What did other patients experience when they reduced their opioid dose?

[Intervention group only]

After watching this video, we will send short text messages (SMS) to your mobile phone. We will send you two SMS every day for 12 weeks. The messages will usually be sent between 9 am and 5 pm. These messages will provide you further information, education, and support. Most people with chronic pain have found these messages useful and supportive while reducing their opioid dose.

These messages are one-way and you are not required to reply. If you want us to stop sending you daily text messages, you can inform us at any time during the study by sending us an email at or by phone [to mention the dedicated phone number] during office hours.

Would you like to receive these daily text messages?

YES / NO

This digital health support is designed by a team of clinicians and researchers who are experienced in pain management together with people with chronic pain who have experienced reducing opioid medications.

We will also send you a text message every 4 weeks. The text message has a link to a short survey to complete online. We will also send the survey link to your email. You can complete the survey using a computer or tablet if you like. We will send you these surveys for 12 weeks.

**We strongly advise you NOT to watch the video, read any messages, or respond to any phone calls while driving or crossing roads to ensure your safety and abidance with the State Government laws.**

**How to contact us?**

You can send us emails or contact us by phone [to mention the dedicated phone number] during office hours if you have questions about the conduct of this study such as completing study forms or using digital support. We will not be always checking or responding to emails or phone calls immediately. Please do not use this in emergencies. The other study phone number [to mention the phone number being used by the SMS software] cannot receive text messages or calls.

If you want to withdraw from the study, you can do so at any time during the study by contacting us via email at or by phone [to mention the dedicated phone number] during office hours.

If you have read this instructions, please click the submit button.

**SUBMIT**
